# First comparison of conventional activated sludge versus root-zone treatment for SARS-CoV-2 RNA removal from wastewaters: statistical and temporal significance

**DOI:** 10.1101/2021.05.09.21256898

**Authors:** Manish Kumar, Keisuke Kuroda, Madhvi Joshi, Prosun Bhattacharya, Damia Barcelo

**Affiliations:** Discipline of Earth Science, Indian Institute of Technology Gandhinagar, Gujarat 382 355, India; Kiran C Patel Centre for Sustainable Development, Indian Institute of Technology Gandhinagar, Gujarat, India; Department of Environmental and Civil Engineering, Toyama Prefectural University, Imizu 939 0398, Japan; Gujarat Biotechnology Research Centre (GBRC), Sector- 11, Gandhinagar, Gujarat 382 011, India; Department of Sustainable Development, Environmental Science and Engineering, KTH Royal Institute of Technology, SE-10044 Stockholm, Sweden; Institute of Environmental Assessment and Water Research (IDAEA-CSIC), Barcelona, and Catalan Institute for Water Research (ICRA)-CERCA, Girona, Spain

**Keywords:** SARS-CoV-2, COVID-19, Environmental Surveillance, Conventional activated sludge process, Root-zone treatment, Wastewater based epidemiology

## Abstract

In the initial pandemic phase, effluents from wastewater treatment facilities were reported mostly free from Severe Acute Respiratory Coronavirus 2 (SARS-CoV-2) RNA, and thus conventional wastewater treatments were generally considered effective. However, there is a lack of first-hand data on i) comparative efficacy of various treatment processes for SARS-CoV-2 RNA removal; and ii) temporal variations in the removal efficacy of a given treatment process in the backdrop of active COVID-19 cases. This work provides a comparative account of the removal efficacy of conventional activated sludge (CAS) and root zone treatments (RZT) based on weekly wastewater surveillance data, consisting of forty-four samples, during a two-month period. The average genome concentration was higher in the inlets of CAS-based wastewater treatment plant in the Sargasan ward (1.25 x 10^3^ copies/ L), than that of RZT plant (7.07 x 10^2^ copies/ L) in an academic institution campus of Gandhinagar, Gujarat, India. ORF 1ab and S genes appeared to be more sensitive to treatment i.e., significantly reduced (p <0.05) than N genes (p>0.05). CAS treatment exhibited better RNA removal efficacy (*p*=0.014) than RZT (*p*=0.032). Multivariate analyses suggested that the effective genome concentration should be calculated based on the presence/absence of multiple genes. The present study stresses that treated effluents are not always free from SARS-CoV-2 RNA, and the removal efficacy of a given WWTPs is prone to exhibit temporal variability owing to variations in active COVID-19 cases in the vicinity and genetic material accumulation over time. Disinfection seems less effective than the adsorption and coagulation processes for SARS-CoV-2 removal. Results stress the need for further research on mechanistic insight on SARS-CoV-2 removal through various treatment processes taking solid-liquid partitioning into account.

**Graphical Abstract:** 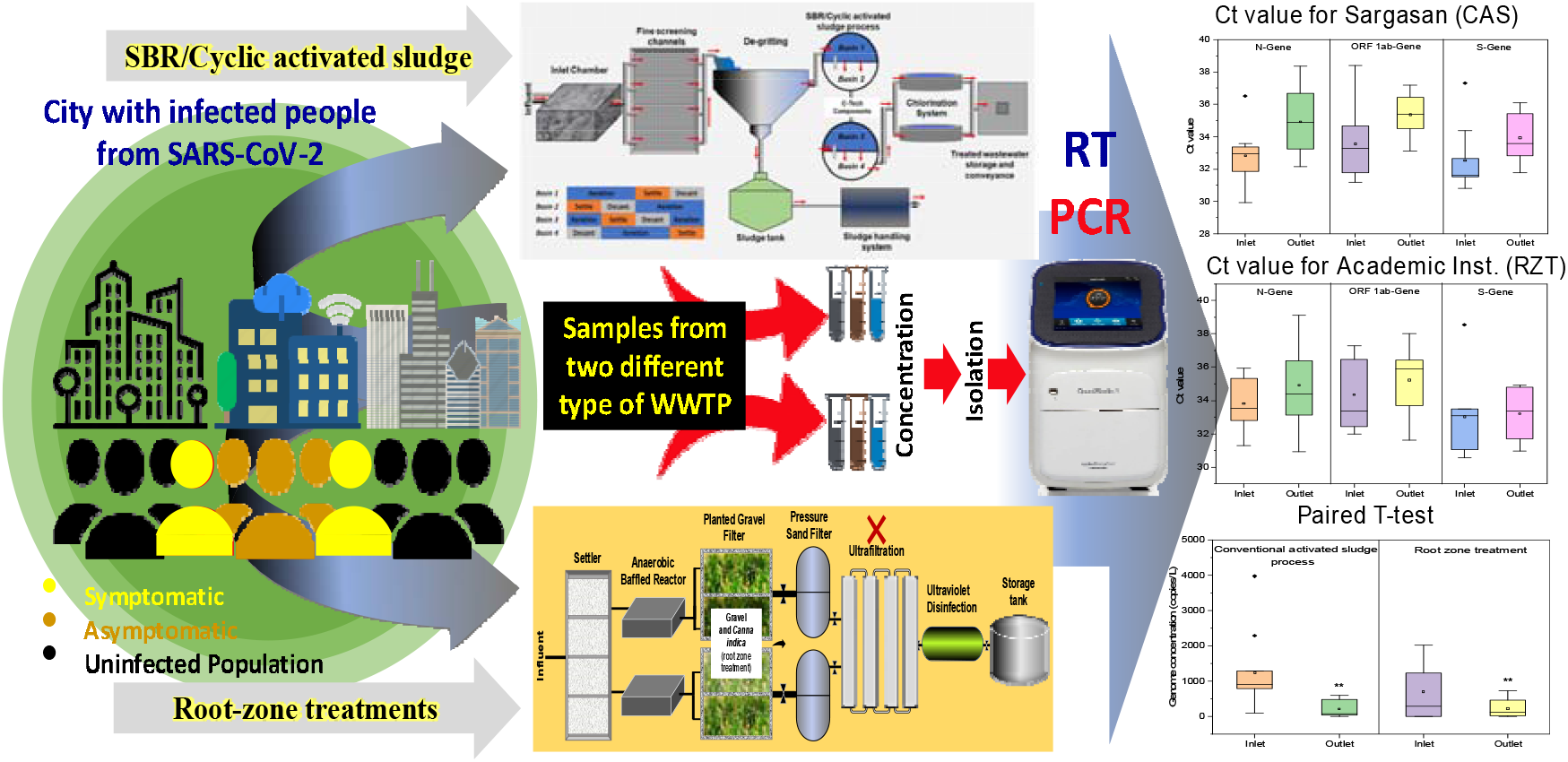

**Highlights:**

- Wastewater treatments may not completely remove the SARS-CoV-2 RNA.
- The activated sludge process exhibited better RNA removal efficacy than root-zone treatment.
- ORF 1ab and S genes appeared more sensitive to treatment than N genes.
- Temporal variability is observed in the removal efficacy of wastewater treatment plants.

## 1. Introduction

At this juncture, when the world is facing a second winter after being threatened for the entire year with Corona Virus Disease (COVID)-19, cases are surging, with over 40 million infections and more than 1 million deaths [1]. To date, we have gained knowledge on many aspects of Severe Acute Respiratory Coronavirus 2 (SARS-CoV-2), especially on transmission, monitoring, analytical techniques, prognosis, diagnosis, models, and management aspects [2-21]. However, the infectivity of SARS-CoV-2 RNA in wastewater, owing to viral shedding of infected symptomatic/asymptomatic patients, and their transmission remains under debate [**22]**. Potential community transmission associated with untreated/treated wastewater, e.g., reuse of wastewater (inbuilt environments), aerosols of wastewater potentially exposing WWTP workers, sludge transfer activities, irrigation and recreational activities in wastewater-impacted waters, is still being debated [23-26]. The two main obstacles are i) whether the viral genome load in wastewater is viable, and ii) whether wastewater treatments can completely remove SARS-CoV-2 RNA? [27-38].

In general, wastewater surveillance of SARS-CoV-2 has focused on early-warning capability verifications [8, 11, 16, 39-40]), or protocol improvement through comparing various techniques of concentration and precipitations [40-43], and solid-aqueous interactions from sludge and virus interaction perspectives. However, since the beginning, subtle parallel efforts were there to check the SARS-CoV-2 RNA presence in secondary- and tertiary-treated wastewater. Apart from several reports neglecting the presence of SARS-CoV-2 in treated water, Randazzo et al., 2020 confirmed 11% (2 out of 18) of secondary- and 0% (0/12) tertiary-treated water samples positive for SARS-CoV-2 RNA. Haramoto et al., (2020) detected as many as 2400 gene copies/L of SARS-CoV-2 RNA in secondary-treated wastewater, whereas raw wastewater samples were not positive with SARS-CoV-2, owing to the difference of sample amounts taken for filtration i.e. 200 mL for raw wastewater vs 5000 mL for secondary-treated wastewater. They also tested river samples, but no positive samples could be traced. Interestingly, they reported that 20% of secondary-treated wastewater samples that were found positive could not show the presence of S and ORF1a genes but the N-genes.

By 2021, more efforts started pouring, which tried to screen the treated water like Westhaus et al., [44] reported modest SARS-CoV-2 removal from all three monitored conventional activated-sludge-based WWTP plants. They pointed out that the plant with full-scale ozonation illustrated a relatively better reduction of SARS-CoV-2 fragments in the effluent; and recommended to include membrane-based WWTP plant for future studies. On the other hand, Hasan et al., [45] reported no positive results after monitoring 11 WWTPs effluents. They concluded that the treatment technologies used in the UAE were efficient in degrading SARS-CoV-2, and confirming the safety of treated water in the country for reuse. Similar results were reported by Balboa et al [27] after observing WWTP in Spain for few days in both effluent and treated sludge.

We previously compared the decay in genetic loading of conventional and Upflow Anaerobic Sludge Blanket (UASB) treatment systems with limited data [13] and reported a gradual decay in gene copies of SARS-CoV-2 from raw influent to UASB effluent to aeration pond and to the final effluents. We then summarized that higher RNA loading translated to higher decay along with the treatment. However, data were based on two-time sampling, and a detailed investigation was recommended. It is still unclear how a varying genome loading in the influent impacts the remaining SARS-CoV-2 genome in the effluent. Therefore, it is novel to perform a comparative study, including both untreated and treated wastewater samples to assess the efficacy of treatment plants. While multivariate analysis (MVA) helps source apportionment for environmental samples, it projects unbiased relationships among parameters and their contribution to variations in the data set [39]. To date, however, reported wastewater surveillance datasets have not been large enough for MVA.

Accordingly, we performed two months of monitoring for SARS-CoV-2 genes in untreated and treated wastewater samples, collected from two mechanically different treatment plants, viz. conventional activated sludge (CAS) process (Sargasan) and root zone treatment (RZT) (academic institution) located in Gandhinagar, India. Our main objectives were to: i) compare and evaluate the removal efficacy of SARS-CoV-2 by CAS and RZT processes through months-long influent and effluent monitoring; and ii) study temporal variations in the removal efficacy of a given treatment process in the backdrop of active COVID-19 cases. We wish to add significant pertinent knowledge related to the actual and varying capabilities of one conventional and another zero-discharge trending root-zone treatment systems, so that infectivity can be adequately understood and appropriate information disseminated to the community. Our study is vital as transmission routes in the developing countries are many, owing to less prevalent, unproperly managed sewer systems that lead to wastewater leakages, occurrences of open defecation and common sewer overflow (CSO) situations.

## 2. Material and Methods

### 2.1 Wastewater treatment plants (WWTPs)

We investigated wastewater samples collected from conventional activated sludge (CAS) based treatment plant situated at the Sargasan ward of Gandhinagar (Sargassan WWTP), and from the root-zone treatment plant of an academic institution located in Gandhinagar, both located in Gujarat, India. Schematic diagrams of the two treatment processes are shown in **Fig. 1**. At Sargasan WWTP (capacity: 10,000 m^3^ /day), the primary treatment consisted of screening by fine screening channels and grit separator tank. The secondary treatment employed was a cyclic activated sludge process operated with 3-5 h, following which the supernatant was removed from the basin and chlorinated to release as the effluent.

**Fig. 1.**
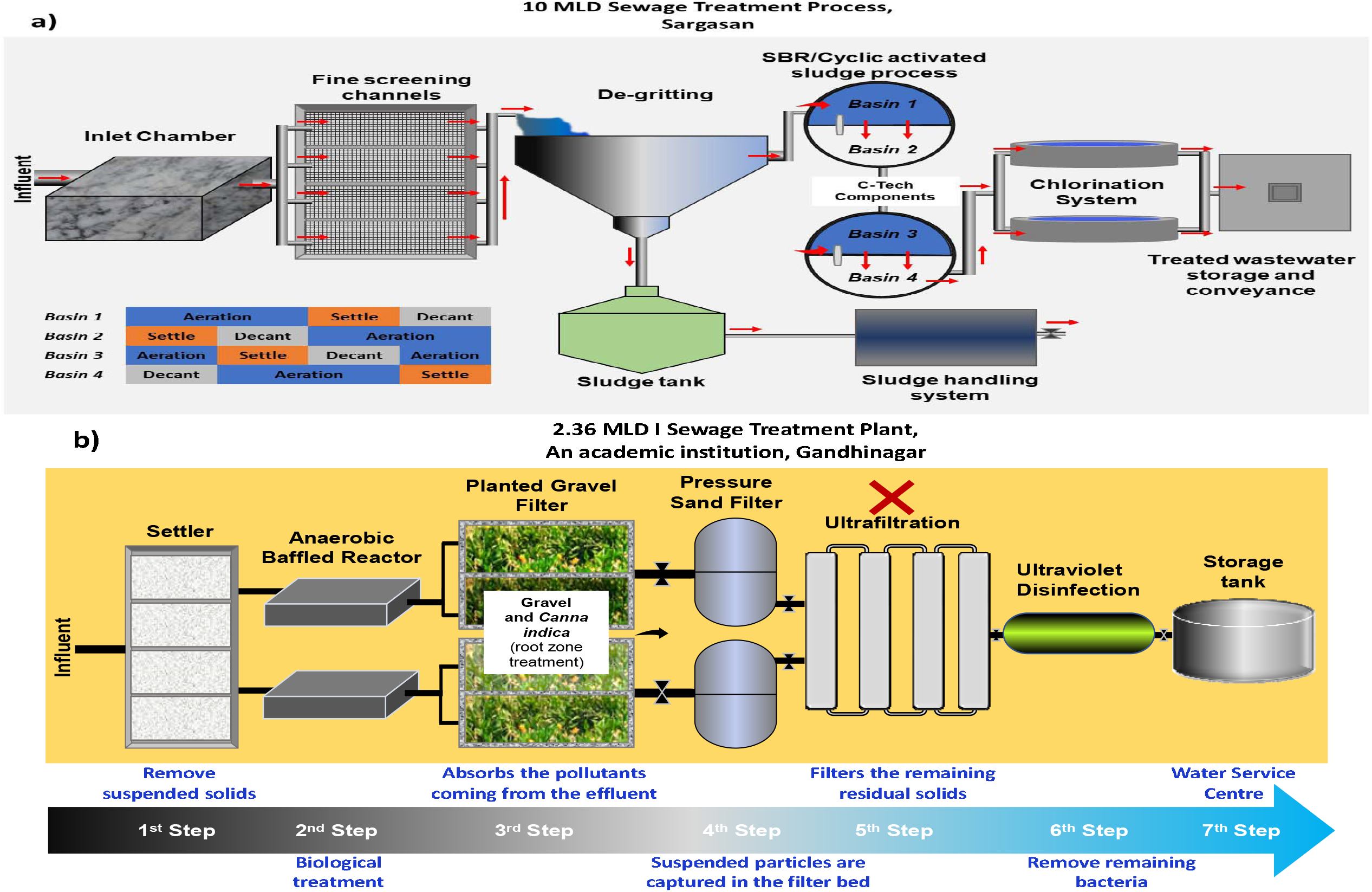
Simplified illustration of the layout of two wastewater treatment plants; a) Conventional Activated Sludge based WWTP in Sargasan, and b) root-zone treatment in an academic institution of Gandhinagar, India monitored during August and September, 2020.

At the treatment plant at the academic institution (capacity: 2,360 m^3^ /day), the root-zone treatment (RZT) was employed as a part of an innovative Decentralized Wastewater Treatment System (DEWATS) that treats all wastewater produced by academic campus dwellers. In this plant, heavy particles and suspended solids in untreated sewage were first removed in the settler tank. Then the sewage was treated by biological treatment through the anaerobic baffled reactor, where anaerobic degradation of organic matter took place. In the third step, the sewage ran through a planted gravel filter, known as an RZT system, where the roots of the *Canna indica* absorbed organic pollutants from the sewage. In the fourth stage, sewage was passed through a pressure sand filter to reduce turbidity and BOD of the effluent. After chlorination, the final effluent was pumped to Water Service Centres in separate storage tanks. Currently, the water does not go through ultrafiltration as it is pumped directly to irrigation tanks to be used for campus irrigation.

### 2.2 Sampling

At the two WWTPs, influent and effluent wastewater samples were initially collected biweekly, then weekly for two months, from August to September 2020. Twenty-one grab samples, representing the treatment plant inlets and outlets of both treatment plants, were collected every Monday of the week at 10 am and placed into 250-ml sterile bottles (Tarsons, PP Autoclavable, Wide Mouth Bottle, Cat No. 582240, India). Simultaneously, blanks were included to check for contamination during travel. The samples were kept cool in an ice-box until analysis. All laboratory analyses were performed on the same day and included duplicates to ensure accuracy and precision. It is imperative to note that we evaluated the removal of SARS-CoV-2 RNA by wastewater treatment methods, including disinfection. It is, therefore, final effluent was sampled after the disinfection process, which is essential in the context of risk assessment of SARS-CoV-2 in receiving water [46].

### 2.3 Detection and extraction of viral RNA from sewage samples

#### 2.3.1 Precipitation of virus

Thirty mL samples were centrifuged at 4000×g for 40 minutes in a 50 mL falcon tube followed by filtration of supernatant using 0.22-micron syringe filter (Mixed cellulose esters syringe filter, Himedia). After filtration, 25 mL of the supernatant was treated with polyethylene glycol (PEG) and NaCl at 80 g/L and 17.5 g/L, respectively and incubated at 10^°^C, 100 rpm overnight. The next day, the mixture was centrifuged for 90 min at 14000×g and the supernatant were discarded to collect a pellet containing viruses and their fragmented genes. The pellet was resuspended in 300µL RNase-free water and kept in 1.5ml Eppendorf tubes at −40 ^°^C, until further analyses.

Briefly, two mechanisms of precipitation are mediated by PEG, which is a chemically inert, nontoxic, water soluble synthetic polymer. a) PEG sterically excludes proteins from a solvent due to ‘salting out effect’ by acting as an “inert solvent sponge”. And b) unfavorable thermodynamic effect on the protein surface charges by solubilized PEG, causing it to be excluded from the “protein zone”, at appropriately high concentrations of polymer. The dynamics of this process is dependent on factors like protein size, their concentration and charge; pH and ionic strength of the solution; and temperature. The required amount of salt depends on the molecular weight of PEG, which counteracts the “Donnan effect” and distributes viruses unequally between the phases.

#### 2.3.2 RNA isolation, RT-PCR and gene copy estimation

A NucleoSpin^®^ RNA Virus, (Macherey-Nagel GmbH & Co. KG, Germany) kit was used for RNA isolation from the pellet containing the concentrated virus. MS2 phage, provided by TaqPathTM Covid-19 RT-PCR Kit, was used as an internal control. Other specifics: a) the nucleic acid was extracted using NucleoSpin^®^ RNA Virus Kit (Applied Biosystems), and Qubit 4 Fluorometer (Invitrogen) was used for RNA concentrations estimation; b) molecular process inhibition control was evaluated through the MS2 phage for QC/QA analyses of nucleic acid extraction and PCR inhibition [47]. We have described methodologies elsewhere [12,13]. Briefly, steps were carried out as per the guideline provided with the product manual of Macherey-Nagel GmbH & Co. KG and RNAs were detected using real-time PCR (RT-PCR).

An Applied Biosystems 7500 Fast Dx Real-Time PCR Instrument (version 2.19 software) was used for SARS-CoV-2 gene detection. A template of 7 µl of extracted RNA was used in each reaction with TaqPath^™^ 1-Step Multiplex Master Mix (Thermofischer Scientific, USA). The reaction mixture volume of 20 µL contained 10.50 µL Nuclease-free Water, 6.25 µL Master Mix, and 1.25 µL COVID-19 Real Time PCR Assay Multiplex. Three controls were included: positive control (TaqPath^™^ COVID-19 Control); negative control (from extraction run spiked with MS2); and a no template control (NTC) [48]. The real-time PCR contained 1 incubation step cycle of 25^°^C for 2 min, 1 cycle of reverse transcription 53^°^C for 10 min, 1 cycle of activation 95^°^C for 2 min, and 40 cycles of amplification, including denaturation at 95^°^C for 3 sec and extension at 60^°^C for 30 sec. Finally, results were interpreted using Applied Biosystems Interpretive Software, and Ct values for three target genes, i.e., ORF1ab, N Protein, and S Protein of SARS-CoV-2, were detected along with MS2 as an internal control.

The samples were considered as positive if at least two genes showed amplification. The average Ct-value of a given sample was then converted to gene copy numbers considering the equivalence of 500 copies of SARS-CoV-2 genes as 26 Ct-value (provided with the kit). The same was extrapolated to derive approximate copies of each gene, using the well-established principle of 3.3 CT change corresponding to a 10-fold gene concentration change. The average effective genome concentration of SARS-CoV-2 present in a given sample was calculated by multiplying the RNA amount used as a template with the enrichment factor for each sample.

It is noteworthy that the primer efficiency of different genes will be slightly varied according to the sequence of primer. However, the gene copies were numbered based on the positive control provided with kit i.e., 10^4^ copies/µl and the final concentration of 25 copies per reaction. Based on several hundreds of RTPCR run, it was found that the positive control was robust enough to provide the same Ct values for all 3 genes, implying no evident difference between the primer efficiency. We report both primary Ct-values and derived gene copies relative to the Ct values of positive controls, for both individual genes and effective SARS-CoV-2 genome concentration.

Due to various constraints, samples were analyzed in duplicate, considering that the samples were analyzed in the batch accompanied with negative and positive controls, and each sample was spiked with known concentrations of MS2. In the event of any variations (among duplicate and controls) of more than 10%, samples were re-analyzed.

### 2.4 Statistical analysis

Box plots were prepared to explain the data variability, and one-way ANOVA was used to determine the significance of the difference among the treatment plant, various gene types and temporal variation in the SARS-CoV-2 RNA copies before and after treatment. The results obtained from ANOVA analysis were reported as (F_critical_= F_calculated_, significant level P) and if F_calculated_ value is greater than F_critical_ value, the null hypothesis will be rejected. The Statistical Package for the Social Sciences (SPSS 21) was used for hypothesis testing and multivariate analyses (MVA) to determine the significance of removal efficacy and relatedness of various water quality parameters with SARS-CoV-2 genes through paired t-tests and principal component analyses (PCA) respectively, after Z-score data normalization [39]. A non-related principal components (PCs) was generated using orthogonal varimax rotation, and the results were projected on three-dimensional loading domain. Since the principal component analysis (PCA) are found to be useful for evidencing temporal variation caused by COVID-19 patient load and treatment, strong positive or negative correlation between a variable and a factor is indicated by a high factor loading close to 1 or −1, respectively. Three-dimensional projection of PCs is an unsupervised pattern recognition technique that groups the objects (variables) as per their similarities within a class and dissimilarities between different classes. In the present study, PCA was done using agglomeration and Ward linkage techniques.

## 3. Results

We analyzed the efficacy of two treatment processes of CAS and RZT (schematic diagrams of the operating mechanism of both plants in Sargasan and academic campus are shown in **Fig. 1 a and b**, respectively). **Table 1** summarizes the change in the Ct-value and gene copies of SARS-CoV-2 N-genes (nucleocapsid protein), S-genes (spike glycoprotein), and ORF 1ab genes (polyprotein) before and after the treatment i.e., in the samples of influent and effluent for two months (August and September 2020) of monitoring. It also provides the date of sampling, effective genome concentration, and active COVID-cases. The Ct values of internal control (MS2 bacteriophage) ranged between 25.41 to 28.01 and 25.59 to 30.08 in the samples from Sargasan and academic institution WWTPs, respectively. No SARS-CoV-2 genes were detected in the negative control samples.

**Table 1.**
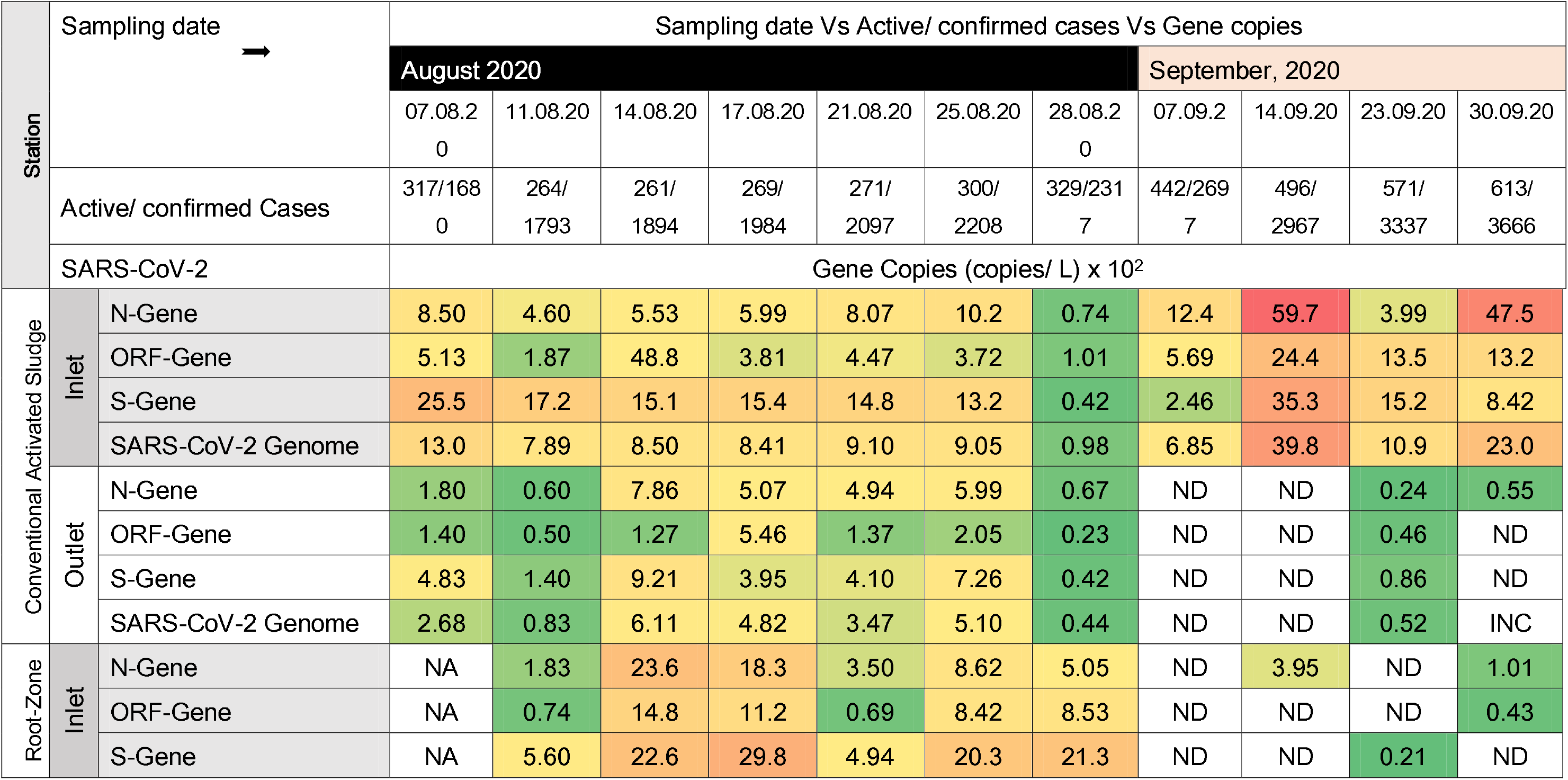
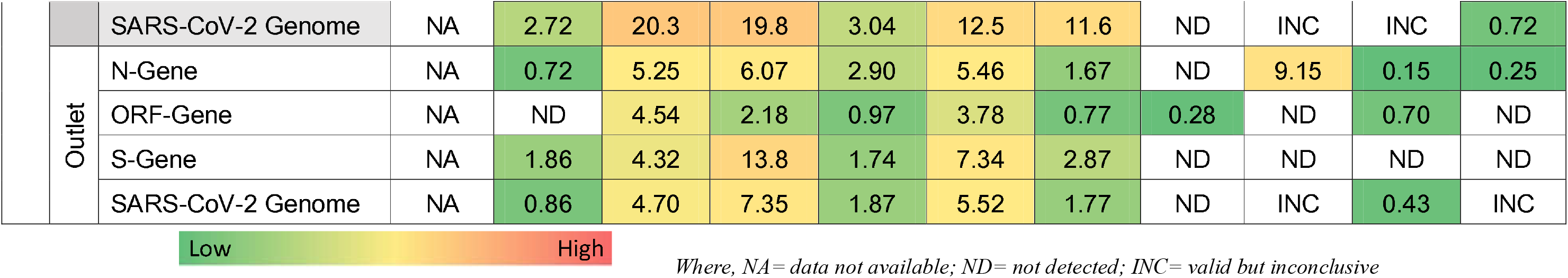
Temporal variation in SARS-CoV-2 genetic material loading found in the influent and effluent samples collected from two different wastewater treatment plants i.e. conventional activated sludge (CAS) at Sargasan ward, and root-zone treatment (RZT) at academic institute at Gandhingar.

Paired T-tests between the inlet and outlet wastewater samples, taken on the same days, were performed to understand the significance of the SARS-CoV-2 gene removal efficacy of each treatment process, i.e., CAS process-based treatment at Sargasan **(Fig. 2a)** and RZT at an academic institution in Gandhinagar **(Fig. 2b)**. We then combined the data and conducted paired T-test analyses of the significance of SARS-CoV-2 gene removal efficacy based on Ct-values obtained and various gene copies calculated for CAS **(Figs. 3a and c)** and RZT **(Figs. 3b and d)**, respectively.

**Fig. 2.**
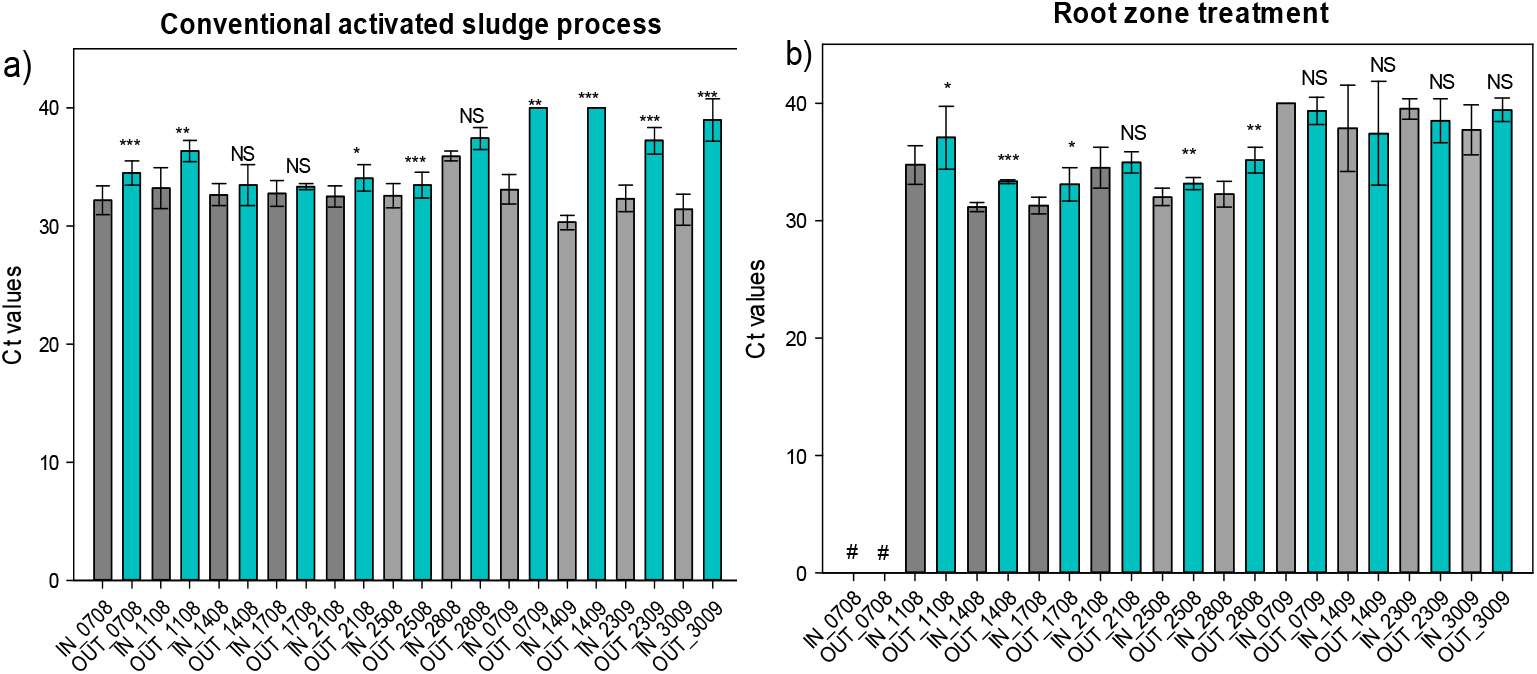
Paired T-test between inlet and outlet wastewater samples taken on the same days for SARS-CoV-2 genetic load in a) Conventional activated sludge process-based treatment at Sargasan, and b) Root-zone treatment at academic institution in Gandhinagar. (where ^***^ = p <0.01; ^**^ = p <0.05; ^*^ = p <0.1; NS = not significant; # = data not available; and RT-PCR was run for 40 cycles).

**Fig. 3.**
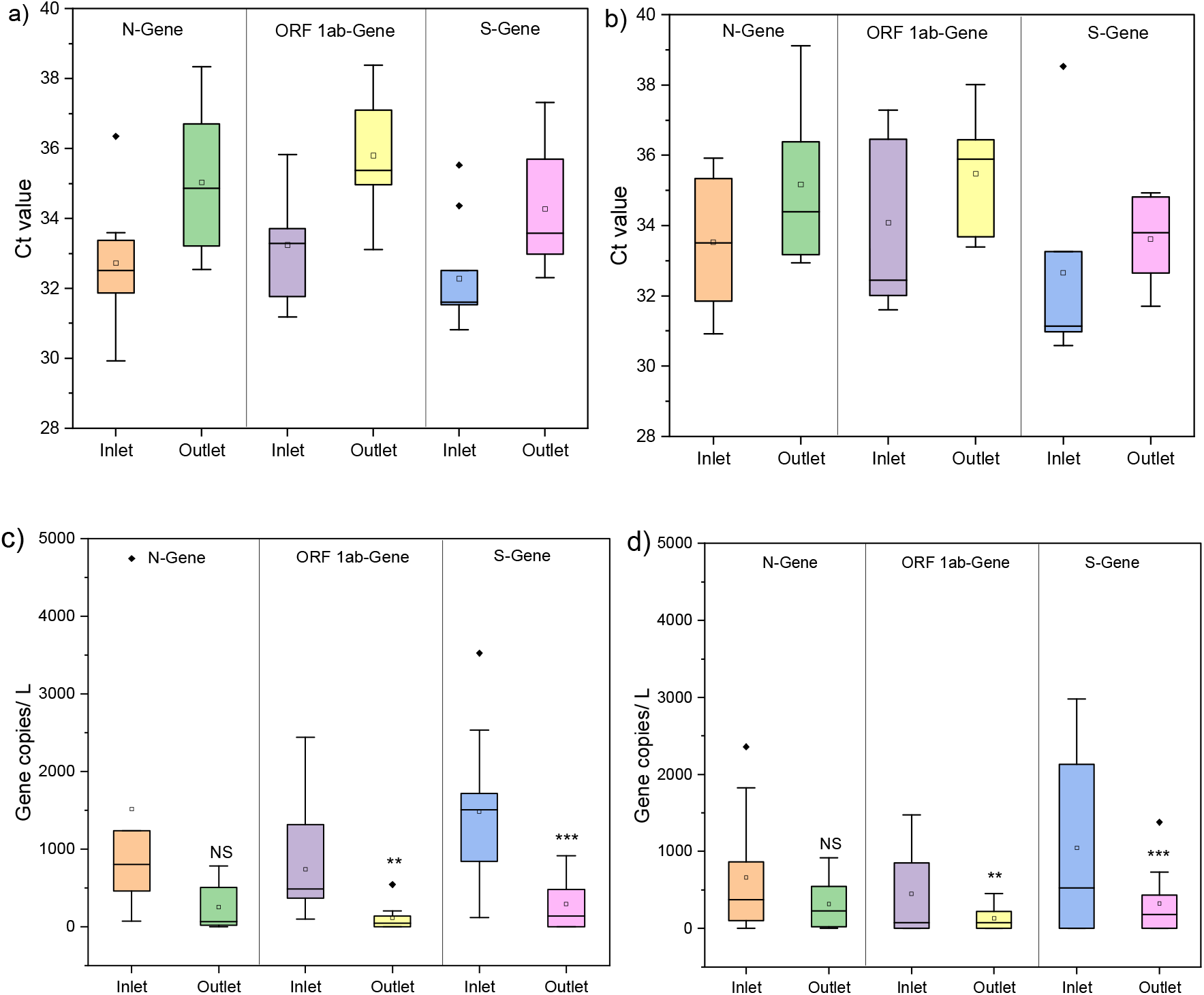
A comparative statistical (paired T-test) analyses of significance of SARS-CoV-2 genes removal efficacy based on Ct-values obtained for a) CAS; and b) RZT; and various gene copies calculated for c) CAS and d) RZT; at p <0.01; p <0.05; and p <0.1 indicated by three, two and one stars. NS signifies not significant.

Overall comparison of SARS-CoV-2 genome removal efficacy of CAS and RZT is expressed through paired T-test performed on the total effective genome concentrations obtained throughout the 60 days of monitoring **(Fig. 4)**. Monthly variations and their significance of SARS-CoV-2 genes removal efficacy of CAS; and RZT is presented in **Fig. 5** to understand the impact of genetic loading in the influent and its correlation with removal efficacy of the treatment processes. MVA was conducted to understand the overall impact of treatment by visualizing the PC loading in a 3-D domain for various water quality parameters and SARS-CoV-2 gene loading of collected influent (untreated) and effluent (treated) samples during the two-month monitoring period (**Figs. 6a and b)**. A summary description of in-situ parameters **(Table S1),** variation explained, eigenvalue variations, and principal component loadings for influent **(Table S2, Fig S1, Table S3)** and effluent **(Table S2, Fig S1, Table S3)** are provided as supplementary material.

**Fig. 4.**
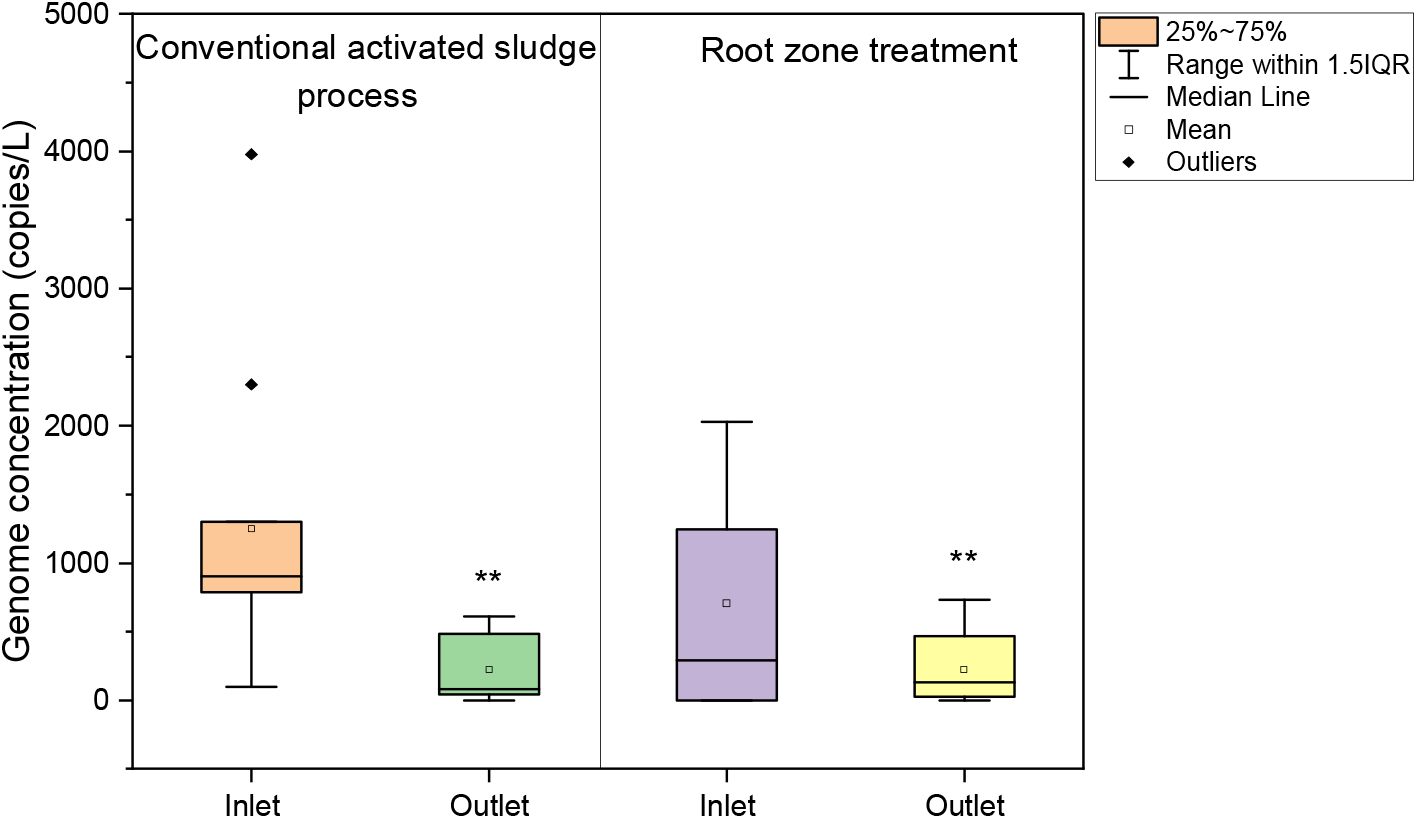
Overall comparison of SARS-CoV-2 genome removal efficacy of conventional activated sludge and root-zone treatments expressed through paired T-test performed on the total effective genome concentrations obtained through out the 60 days of monitoring period. Same level of significance is used as above.

**Fig. 5.**
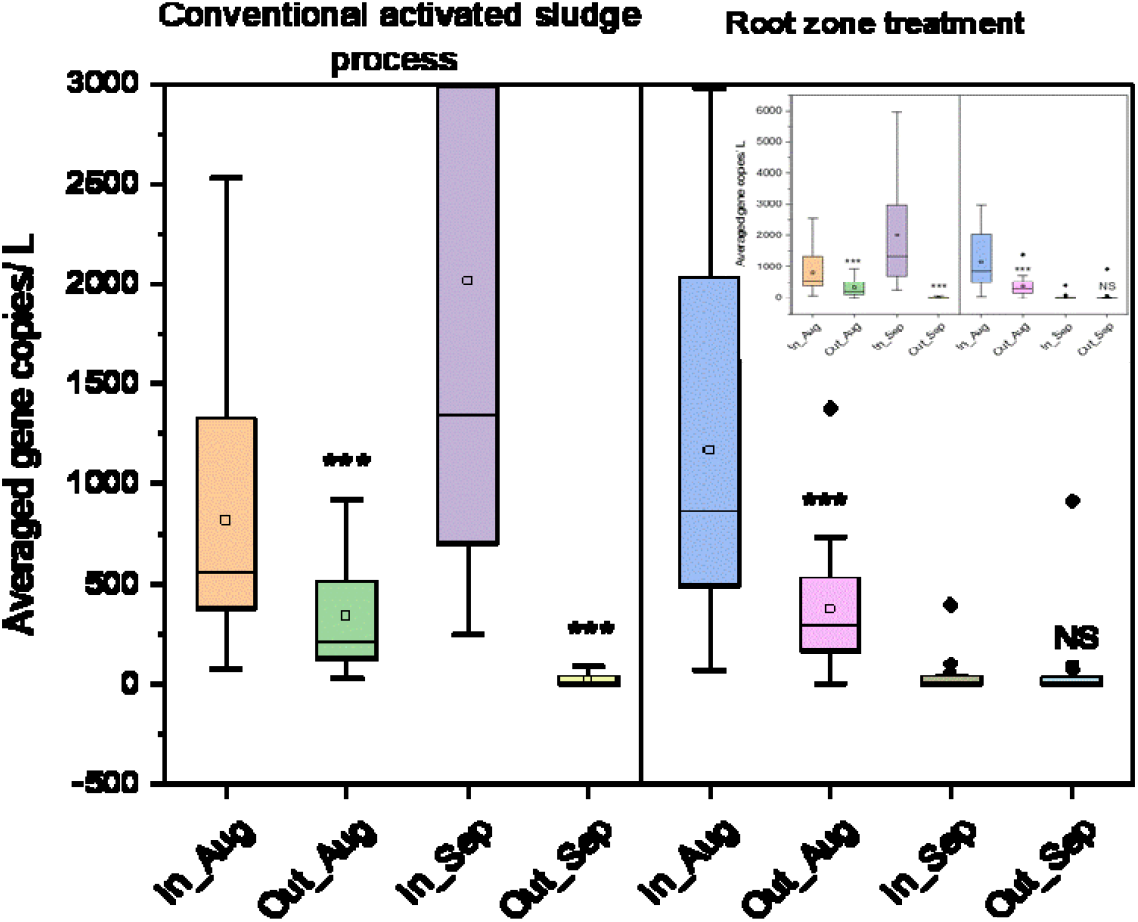
A comparative statistical (paired T-test) analyses in monthly variation of significance of SARS-CoV-2 genes removal efficacy of CAS; and b) RZT; at p <0.01; p <0.05; and p <0.1 indicated by three, two and one stars. NS signifies not significant.

**Fig. 6.**
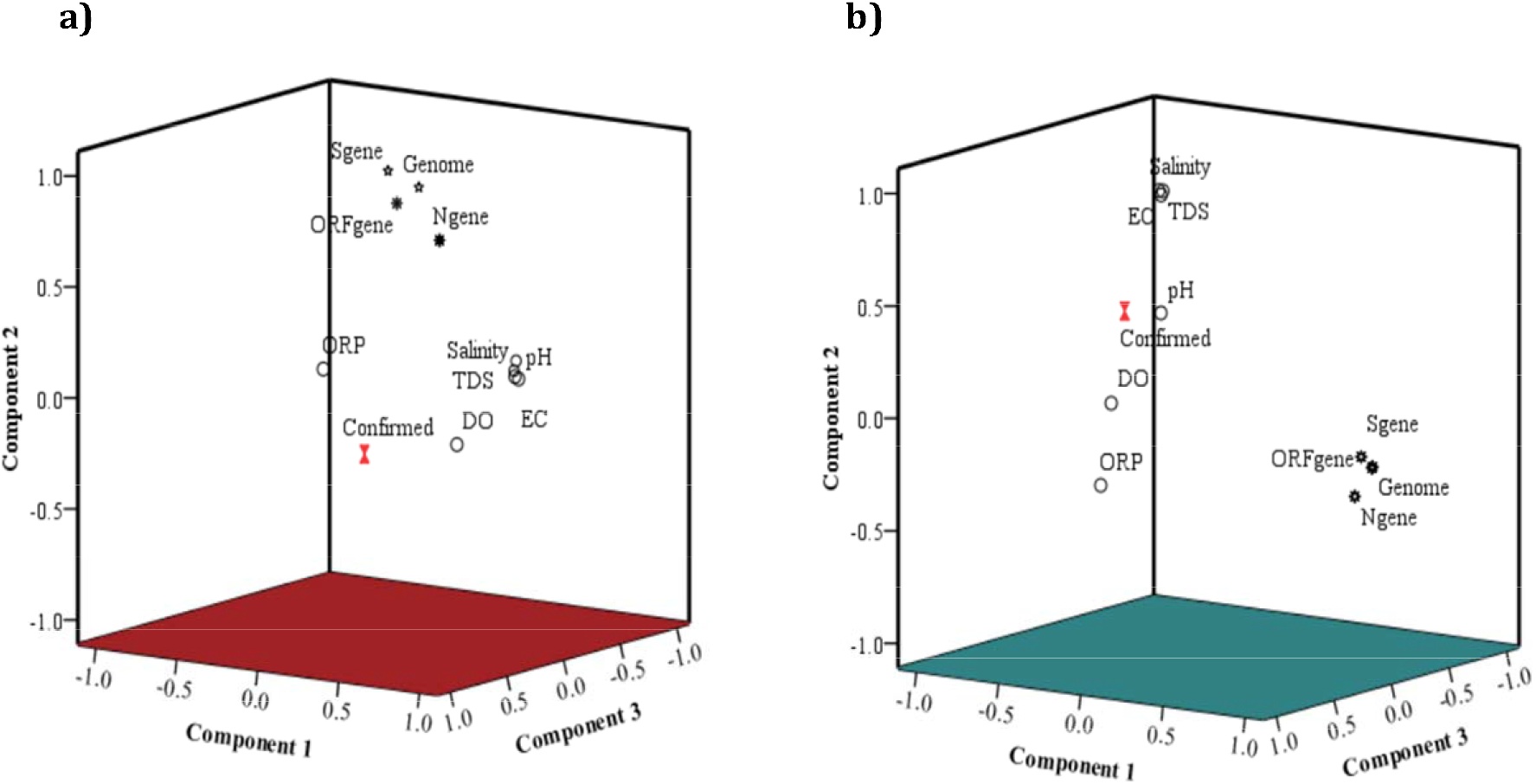
Three-dimensional projection of the principal component loading for a) Influent and b) effluent; exhibiting the effect of treatment on SAR-CoV-2 genes association with other water quality parameters and confirmed cases of COVID-19.

Although there will be a considerable uncertainty, we could estimate the number of people shedding SARS-CoV-2 to wastewater. SARS-CoV-2 is contained in the human stool at 4-6 log copy/g [49], and assuming that the average stool weight is 500 g per day per person, that results in 5×10^6^ to 5×10^8^ copies per person per day shredded to wastewater. Assuming that our raw wastewater samples had 1000 copies/L on average, raw wastewater from Sargassan WWTP had 1×10^9^ copies per day, implying that there were 2 to 200 people shedding SARS-CoV-2 in the catchment on a day. However, there would be too many uncertainties in this calculation, due to significant decay/reduction of viral RNA during transport from toilets to WWTPs. Therefore, hereafter, only Ct-values and gene copies are compared. Further, the role of aqueous and solid-phase interactions for the quantification of SARS-CoV-2 gene concentrations has been prominently highlighted in terms of recovery of the viral RNA in the aqueous environment through solid fractions [50]. However, we did not take sludge into account as there still needs a robust standard protocol for sludge clean-up and RT-qPCR measurements to be established.

## 4. Discussion

### 4.1 Significance of Treatment

Of the eleven samples collected from the inlet and outlet points of WWTPs during the study period, eight samples from Sargasan and five samples from the academic institution showed significant removal of the viral genes **(Figs. 2a and 2b)**. Paired T-tests between influent and effluent wastewater show a significant reduction through CAS treatment systems except for three occasions. Reduction/removal of SARS-CoV-2 genes was highly significant (p <0.01) in nearly 50% of the samples, with non-significant removal in August only. RZT appeared effective in August but failed to show significant removal of SARS-CoV-2 RNA in September. There may be two possible explana tions related to the operation of WWTPs and COVID-19 cases in the vicinity of WWTPs. The RZT was situated and precisely received waste from the campus dwellers and visitors only, and COVID-19 cases increased in September 2020. Thus, even if we assume the viral shedding contribution of visitors was non-variable, it is certain that genetic loading increased in the RZT plant during September 2020. We also suspect that operating conditions at the treatment plants were not consistent throughout the monitoring period. Nevertheless, the RZT achieved significant removal on more than 50% of the sampling dates.

Paired t-tests show that irrespective of treatment type, the N-gene is much more stable? than S- and ORF-1ab genes of SARS-CoV-2 **(Figs. 3a to d)**. Removal efficacy was highest for S-genes (p <0.01) followed by ORF-1ab (p <0.05) for both treatment processes. Overall, N-genes showed non-significant reduction after treatment. The ORF 1ab-gene copy numbers decreased by 84.4% (*t*=2.78, *p*=0.022) and 70.5% (*t*=2.30, *p*=0.047) in Sargasan WWTP and the academic institution WWTP, respectively **(Fig. 3c and d)**. Likewise, S-genes were significantly removed by both treatment plants (80.5%, *t*=4.10, *p*=0.002 at Sargasan and 69.5%, *t*=2.84, *p*=0.019 at the academic institution). Conversely, the abundance of N-gene declined 83.4% at Sargasan WWTP **(Fig. 3c)** and 52.0% at the academic institution during treatment **(Fig. 3d)**, but the differences in S- and N-gene removal were statistically significant (*t*=2.04, *p*=0.069 and *t*=1.59, *p*=0.147, respectively). The results showed that both the cyclic activated sludge process and root zone treatment plants of Sargasan and the academic institution effectively removed ORF ab-genes and S-genes, but not N-genes.

Our hypothesis-prevalence may be causing the difference in removal- was not correct **(Table 1)**. It seems structural properties of the genes are more responsible for such removal disparity than prevalence. This is because, among four major structural proteins of SARS-CoV2; S proteins are the most exposed one being the spike surface glycoprotein (S), while ORF-1ab gene is not only a signatory gene for SARS-CoV-2 genes but also located at both the 5’ & 3⍰-terminuses of the SARS-CoV-2 genome [37]. Nucleocapsid protein (N) is more protected in the SARS-CoV-2 structures, and common genes among family *Coronaviridae*, marked by the presence of single-stranded, positive-sense RNA genome, surrounded by spikes and protein envelope.

A comparison of the effectiveness of various wastewater treatment systems for the removal of SARS-CoV-2 genetic material is shown in **Table 2**. Earlier studies suggested reduction of SARS-CoV-2 genetic material during wastewater treatment processes via secondary treatment such as activated sludge/ A2O/ extended aeration and tertiary treatment such as disinfection, coagulation, flocculation, sand filtration, NaClO/UV [21]. Interestingly, none of the studies investigated the removal efficacy of a given treatment for SARS-CoV-2 RNA. In our study, both the CAS and RZT processes are found to effectively remove SARS-CoV-2 RNA. To the best of our knowledge, this is the first report assessing the effectiveness of RZT for SARS-CoV-2 RNA reduction.

**Table 2.** Comparison of the effectiveness of various wastewater treatment systems for the removal of SARS-CoV-2 genetic material

| Country | City | Wastewater treatment method and types | Virus concentration method | RT-(q)PCR target region | Before treatment (gene copies /L) | After treatment (gene copies /L) | References |
| --- | --- | --- | --- | --- | --- | --- | --- |
| India | Gandhinagar | Root Zone Treatment/ institutional wastewater | PEG precipitation | N gene | $6.58 \times 10^2$ | $3.16 \times 10^2$ | **Present study** |
| | | | | ORF 1ab gene | $4.48 \times 10^2$ | $1.32 \times 10^2$ | |
| | | | | S gene | $1.05 \times 10^3$ | $0.32 \times 10^3$ | |
| | | | | Genome conc. | $7.07 \times 10^2$ | $2.27 \times 10^2$ | |
| | | SBR/Cyclic Activated Sludge Process/ chlorination Municipal wastewater | PEG precipitation | N gene | $1.48 \times 10^3$ | $0.25 \times 10^3$ | |
| | | | | ORF 1ab gene | $0.74 \times 10^3$ | $0.12 \times 10^3$ | |
| | | | | S gene | $1.49 \times 10^3$ | $0.29 \times 10^3$ | |
| | | | | Genome conc. | $1.25 \times 10^3$ | $0.22 \times 10^3$ | |
| | Ahmedabad | UASB | PEG precipitation | ORF1ab, N gene<br>S gene | $3.5 \times 10^3$ | <LOQ | [13] |
| | | Aeration pond | | ORF1ab | $1.5 \times 10^2$ (< LOQ) | Not detected | |
| China | | Septic tank treatment of hospital effluent | PEG precipitation | ORF1<br>N gene | Not detected | $0.05\text{--}1.87 \times 10^3$ | [20] |
| France | Paris | Municipal wastewater treatment | Ultracentrifugation | E gene | $1 \times 10^3\text{--}1 \times 10^5$ | $<10 \times 10^3$ | [32] |
| Spain | Murcia | Secondary treatment | Aluminium | N gene | N1: $1.4 \times 10^3$ | $<2.5 \times 10^3$ | [21] |
| | | (activated sludge/ A2O/ extended aeration), disinfection, NaClO/UV) | flocculation – beef extract precipitation | | N2: $3.4 \times 10^3$<br>N3: $3.1 \times 10^3$ | | |
| | Valencia | Municipal wastewater treatment (treatment methods not provided) | Aluminium flocculation – beef extract precipitation | N gene | N1: $1.0 \times 10^3 - 1.0 \times 10^4$<br>(Averaged value) | Not detected | [21] |
| | Ourense | Primary settler, secondary treatment of municipal sewage | Ultrafiltration of centrifugated supernatant | N gene<br>E gene<br>RdRp gene | $7.5 \times 10^3 - 1.5 \times 10^4$ | Not detected | [27] |
| **Australia** | Brisbane | Untreated wastewater | Adsorption-direct RNA extraction and Ultrafiltration | N Sarbeco, NIID_2019-nCOV_N | $1.9 \times 10^1 - 1.2 \times 10^2$ copies/ L | NA | [8] |
| **USA** | Southern Louisiana | Untreated wastewater, secondary treated, and final effluent | Ultrafiltration and Adsorption-elution using electronegative membrane | CDC N1, N2 | $3.1 \times 10^3 - 7.5 \times 10^3$ | Not detected | [40] |
| **Netherlands** | - | Untreated wastewater | Ultrafiltration | CDC N1, N2, N3, E_Sarbeco | $2.6 \times 10^3 - 2.2 \times 10^6$ | NA | [61] |
| **Italy** | Milan and Rome | Untreated wastewater | PEG/dextran precipitation | RT-qPCR (RdRp), | 6/12 samples found positive; | NA | [62] |

|  |  |  |  | nested PCR<br>(ORF1ab and S<br>assays) | gene copies<br>were not<br>detected |  |  |
| --- | --- | --- | --- | --- | --- | --- | --- |
| **Japan** | Yamanashi<br>Prefecture | Untreated influent and<br>secondary-treated<br>wastewater before<br>chlorination | Electronegative<br>membrane-vortex<br>(EMV) method and<br>the membrane<br>adsorption-direct<br>(MAD) RNA<br>extraction method | N_sarbeco,<br>NIID_2019-nCOV_N,<br>CDC-N1, N-2 | EMV: $<6.6 \times 10^4$<br>– $<8.2 \times 10^4$<br>MAD: $<4 \times 10^3$ | EMV: $<1.4 \times 10^2$ – $2.5 \times 10^3$<br>MAD: $<1.6 \times 10^2$ | [29] |
| **USA** | Bozeman,<br>Montana | Untreated wastewater | Ultrafiltration | CDC N1, N2 | $>3 \times 10^4$ | NA | [34] |
| **USA** | Massachusetts | Untreated wastewater | PEG precipitation | CDC N1, N2,<br>N3 | $>2 \times 10^5$ | NA | [63] |
| **France** | Paris | Untreated and treated<br>wastewater | Ultracentrifugation | E_Sarbeco | $>10^{6.5}$ | $\sim 10^5$ | [64] |

### 4.2 Comparative efficacy of CAS and RZT processes to remove SARS-CoV-2 genes

SARS-CoV-2 RNA is substantially reduced in treated wastewater i.e. effluents of both WWTPs throughout the sampling period, as indicated by the overall comparison of SARS-CoV-2 genome removal efficacy of CAS and RZT through a paired T-test **(Fig. 4.).** Although there was a significant difference in average SARS-CoV-2 genome concentration in the influents of the CAS plant at Sargasan (1.25 x 10^3^ copies/ L) and the RZT system of an academic institution (7.07 x 10^2^ copies/ L). Yet, both processes mostly showed effective removal at p<0.05. However, incomplete removal may have some environmental and health implications.

While infectivity and viability of these genomes are still being debated and researched with a general consensus of viability being less likely and thus the infectivity, there is still no study that has yet proven the chance of transmission and infectivity impossible. In such a scenario, significant removal is not enough, as such effluents will finally be received by the ambient waters. Therefore, we foresee an immediate increase in reporting of SARS-CoV-2 genes in freshwater systems like lakes, rivers, and perhaps groundwater. Several imperative hypotheses need to be tested in this regard, and the present study signifies the need of such investigations.

Further, we also suspect that the size of the treatment plant and operational and management consistencies, along with the quality of influent water will play a critical role in the entire research scenario of COVID-19 transmission and monitoring [13]. As far as treatment type is concerned, the RZT will show a bit wider fluctuation than the CAS treatment process **(Fig. 4).** The low genome concentration at the academic institution WWTP is apparently due to institutional wastewater load which was confined to the institutional community and malfunctioning of the ultrafiltration unit of the WWTP. Conversely, the Sargasan WWTP receives municipal wastewater, resulting in the presence of SARS-CoV-2 RNA in effluent wastewater, owing to fluctuating genetic loading in the inlet waters. We conclude that both WWTPs effectively removed viral genes, but Sargasan STP was more efficient (82.4% decrease, *t*=2.98, *p*=0.014) than the academic institution (67.9% decrease, *t*=2.54, *p*=0.032) **(Fig. 4).** It is imperative to note that we have collected samples from both treatment processes after disinfection processes and still found the genetic fragments of SARS-CoV-2 in the effluent. This observation may imply that owing to nano-sized colloidal nature of genetic fragments, disinfection processes like chlorination/UV are likely to be less effective than the process of coagulation.

Overall, as PCR-based detection of RNA does not mean detection of viable SARS-CoV-2, and quantifying active (viable) SARS-CoV-2 is a difficult challenge, with so far only one lab-scale experiment reported (Bivins et al. 2020), we recommend further study for a valid discussion on implications of leftover SARS-CoV-2 RNA after the treatment. However, our data explicitly disapprove the general notion that treatment completely removes the genetic fragments of SARS-CoV-2.

### 4.3 Temporal variation in removal efficacy

As suspected above, we investigated the role of influent quality in terms of SARS-COV-2 genetic loading through temporal variation in the performances of both CAS and RZT systems **(Fig 5).** For CAS plant in Sargasan ward, inlet quality in September showed higher genetic loading than that of August, which has been verified by confirmed COVID-19 cases in the city, yet removal was better in September than August 2020. When inquired with operational staff, it seems that operational inconsistencies are responsible for these results rather than the genetic material loading. While in the case of the academic institution RZT-based plant, where the operation was rather more consistent, it seems that genetic material loading in the inlet water has reflected the genome concentration left in the effluent waters. This is also very likely to be attributed to the size of plant i.e., CAS facility of Saragasan is 10,000 m^3^ /day against 2360 m^3^ /day of the RTZ plant of the academic institution, leading to the sensitivity of RZT plant for genetic loading in the inlet wastewater. Nevertheless, at this juncture, we take these results as indicative ones, and more convincing conclusions pertaining to the role of influent water quality, and its implication may be derived after further monitoring. Such notion has also been expressed elsewhere [51-54].

### 4.4 Treatment Impact Insight through multivariate statistical analyses

Principal component analyses show a comprehensive picture of the overall contribution and influence of treatment on SARS-COV-2 gene removal. The entire dataset obtained for influent and effluent were subjected to PCA and projected in the 3-D domain of three main PCs. Owing to more complex nature of influents, four PCs were identified after nine iterations that explain 90% of the total variance in the dataset of influent waters. The first PC explains 34% of the total variance with significant loading for *in-situ* water quality parameters forming a cluster (EC, TDS, Salinity, and pH) with moderate loading (0.5) for N-genes (**Fig. 6a** and Supplementary **Table S2** and **S3**). On the other hand, nearly the same (∼30%) variation of data sets is explained by SARS-COV-2 genes, and genome concentrations form a cluster upper left domain with significant loadings for effective genome concentrations (0.94) followed by S-genes, ORF-1ab, and N-genes as PC2. Interestingly in influent waters, N-genes illustrated moderate to high loading as both PC1 and PC2.

After treatment, the complexion changed significantly with the overall reductions of PCs to three, explaining cumulative variations of 80% in the dataset. Another significant observation was that SARS-CoV-2 genes exhibit higher loadings than the *in-situ* water quality parameters in effluent waters. Order of loadings among SARS-CoV-2 genes and genome remains the same i.e., effective genome concentration>S-genes>ORF-1ab>N-genes. Confirmed COVID-19 emerged as PC3 (with moderate loading of 0.78) in influent waters, stressing the relationship of confirmed cases with SARS-CoV-2 RNA in the wastewater, but the influence was weakened in the treated water with non-significant say in the quality variations of the samples [55-60].

This is the first time MVAs was used with wastewater surveillance dataset to signify the impact of treatment, which eventually proves that: i) wastewater surveillances did track COVID-19 loading of the community; ii) influent waters present a better picture in terms of SARS-CoV-2 gene monitoring; iii) effective genome concentration should be calculated based on presence/absence of multiple genes rather the presence of one specific gene; iv) N-genes are the most resistant to treatment with higher sensitivity than S and ORF-1ab genes; and v) the presence of residual SARS-CoV-2 genes after treatment is critical from the effluent quality point of view. Among the other exciting observations; the explicit grouping/clustering of SARS-CoV-2 genes and other water quality parameter; and influence of confirmed COVID-19 cases has been significant from the wastewater-based epidemiology perspectives.

## 5. Conclusion

A comparison of SARS-CoV-2 RNA removal efficacy of CAS and RZT, the two most used treatment systems in India, was studied through biweekly and monthly variations in their performances. We applied long-term monitoring data and performed statistical tests to understand the significance of removal and correlated it with other water quality parameters before and after deployed treatment. For the first time, MVAs used in this study along with other statistical tests highlighted the disparity in performance and statistical significance of SARS-CoV-2 RNA removal between CAS and RZT. It can be concluded that influent waters present better picture in terms of SARS-CoV-2 gene monitoring; effective genome concentration should be calculated based on presence/absence of multiple genes rather the presence of one specific gene; and treatments are less effective on N-genes and the most effective for S-genes. CAS treatment exhibited better RNA removal rate (*t*=2.98, *p*=0.014) compared to the root-zone treatment (*t*=2.54, *p*=0.032). In addition, treatment plants with smaller capacity are likely to show more fluctuations in effluent water quality.

Two most critical findings from the ongoing pandemic perspectives were that the treated effluents are not always free from SARS-CoV-2 RNA, and are subject to temporal variability. We stress the need for wastewater surveillance of SARS-CoV-2 at the treatment plant scale with further investigation on the efficacy of the treatment processes on the removal of the enveloped virus such as SARS-CoV-2 as well as the genomic materials. The future research efforts may therefore consider the influence of genetic material loading in the influent, difference in sewage flow and treatment methods, hydraulic and sludge retention time of technology used, and serviced people. In addition, the mechanistic understanding may be generated on the SARS-CoV-2 removal using long-term step-wise sampling and monitoring of a given treatment processes. Nevertheless, our results are based on RNA fragment detection by RT-PCR, thus the abundance of viable SARS-CoV-2 in the samples can be significantly lower than the RNA-based gene copies. Therefore, research is needed for assessing infectivity through viable virus estimation, specifically for the use of reclaimed water in agriculture and drinking water supply.

## Data Availability

All data is included. If necessary additional information will be provided on demand.

## Notes

The authors declare no competing financial interest.

## Acknowledgment

This work is funded by Kiran C Patel Centre for Sustainable Development at IIT Gandhinagar, UNICEF, Gujarat, and UKIERI. We also acknowledge the help received from Dr. Arbind K Patel and other GBRC staff who contributed towards sample and data analyses.

