## Supplements for "First comparison of conventional activated sludge versus root-zone treatment for SARS-CoV-2 RNA removal from wastewaters: statistical and temporal significance"

**Supplementary Information-**

Table S1: In-situ water quality of influent and effluent collected during the monitoring period.

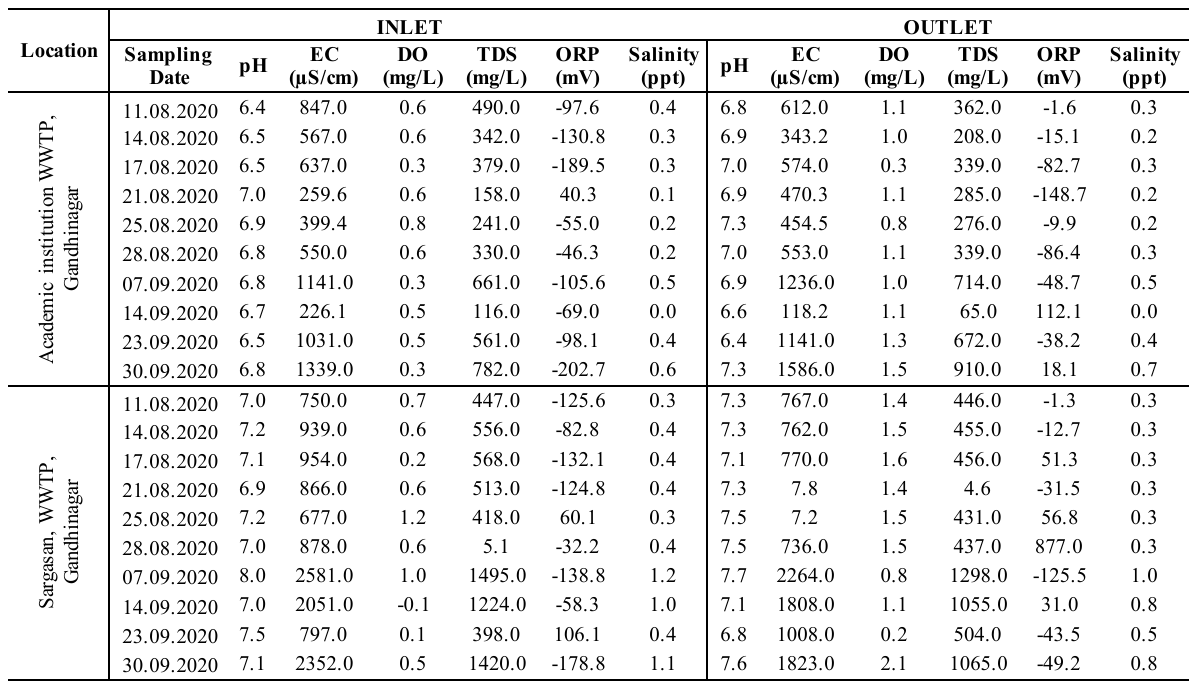

**S-II A: Multivariate results for Influent i.e. untreated samples**

| **Table S2. Total Variance Explained** | | | | | | | | | |
| --- | --- | --- | --- | --- | --- | --- | --- | --- | --- |
| **Component** | **Initial Eigenvalues** | | | **Extraction Sums of Squared Loadings** | | | **Rotation Sums of Squared Loadings** | | |
|  | **Total** | **% of Variance** | **Cumulative %** | **Total** | **% of Variance** | **Cumulative %** | **Total** | **% of Variance** | **Cumulative %** |
| **1** | **5.190** | **47.180** | **47.180** | **5.190** | **47.180** | **47.180** | **3.754** | **34.123** | **34.123** |
| **2** | **2.303** | **20.932** | **68.113** | **2.303** | **20.932** | **68.113** | **3.398** | **30.893** | **65.016** |
| **3** | **1.471** | **13.375** | **81.487** | **1.471** | **13.375** | **81.487** | **1.452** | **13.200** | **78.216** |
| **4** | **1.001** | **9.099** | **90.586** | **1.001** | **9.099** | **90.586** | **1.361** | **12.370** | **90.586** |
| **5** | **.611** | **5.555** | **96.141** |  |  |  |  |  |  |
| **6** | **.249** | **2.266** | **98.407** |  |  |  |  |  |  |
| **7** | **.084** | **.766** | **99.172** |  |  |  |  |  |  |
| **8** | **.052** | **.474** | **99.646** |  |  |  |  |  |  |
| **9** | **.038** | **.346** | **99.992** |  |  |  |  |  |  |
| **10** | **.001** | **.006** | **99.998** |  |  |  |  |  |  |
| **11** | **.000** | **.002** | **100.000** |  |  |  |  |  |  |
| **Extraction Method: Principal Component Analysis.** | | | | | |  |  |  |  |

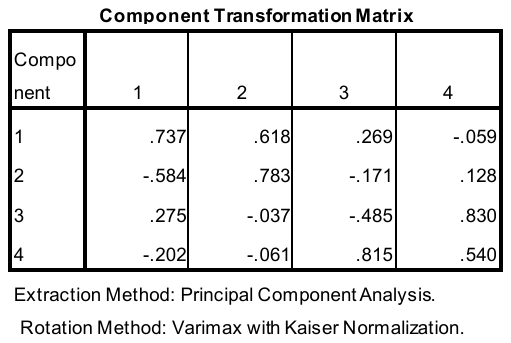

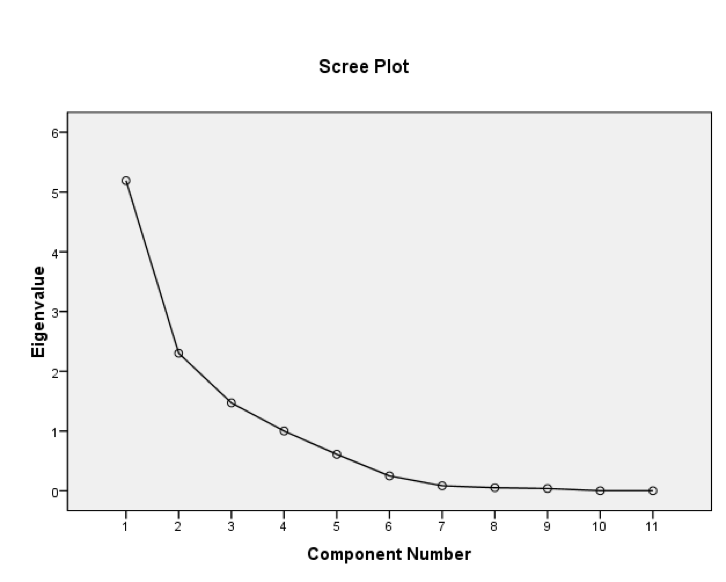
**Figure -S1. Eigen value variation along components and component transformation matrix.**

**Table S3.: Rotated component matrix with loading for four principle components.**

|  | | | | |
| --- | --- | --- | --- | --- |
|  | Component | | | |
|  | 1 | 2 | 3 | 4 |
| Confirmed | .430 | -.144 | .783 | .137 |
| Ngene | .495 | .744 | .213 | -.074 |
| ORFgene | .267 | .894 | .263 | .138 |
| Sgene | -.113 | .935 | -.200 | .004 |
| Genome | .287 | .944 | .099 | -.012 |
| pH | .618 | .055 | -.309 | .639 |
| EC | .947 | .172 | .193 | -.098 |
| DO | -.031 | -.361 | -.692 | .258 |
| TDS | .925 | .234 | .151 | -.145 |
| ORP | -.356 | .049 | .023 | .898 |
| Salinity | .946 | .199 | .199 | -.073 |
| Extraction Method: Principal Component Analysis.  Rotation Method: Varimax with Kaiser Normalization. | | | | |
| a. Rotation converged in 9 iterations. | | | |  |

**S-B: Multivariate results for Influent i.e. untreated samples**

**Table S4: Total Variance Explained**

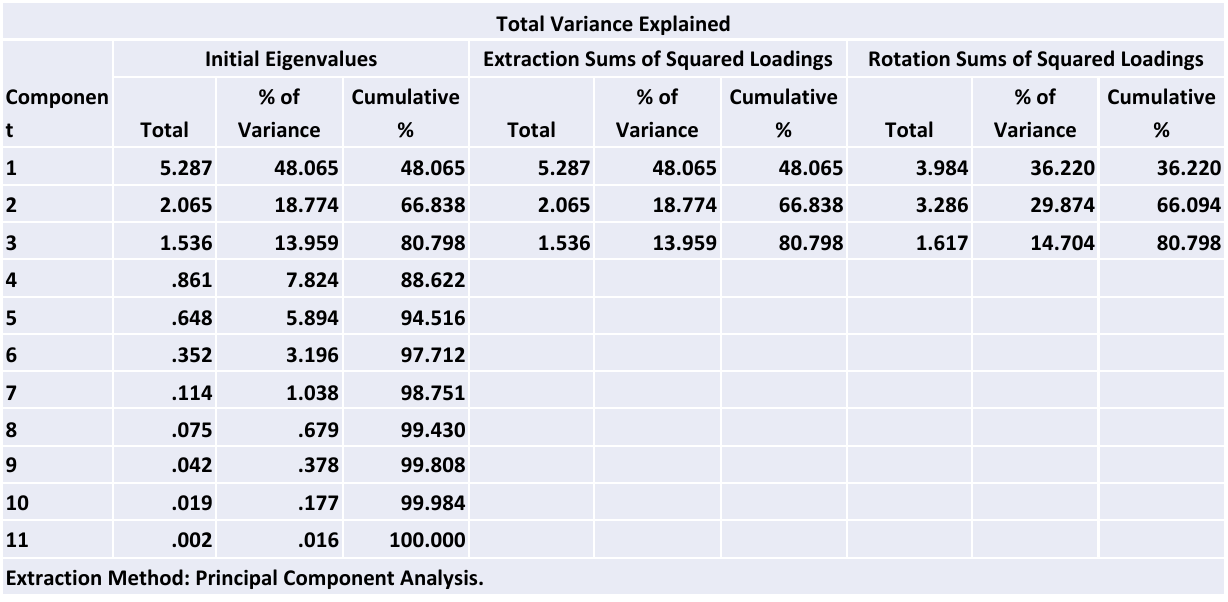

**Figure S2: Eigen value variation along components and component transformation matrix.**

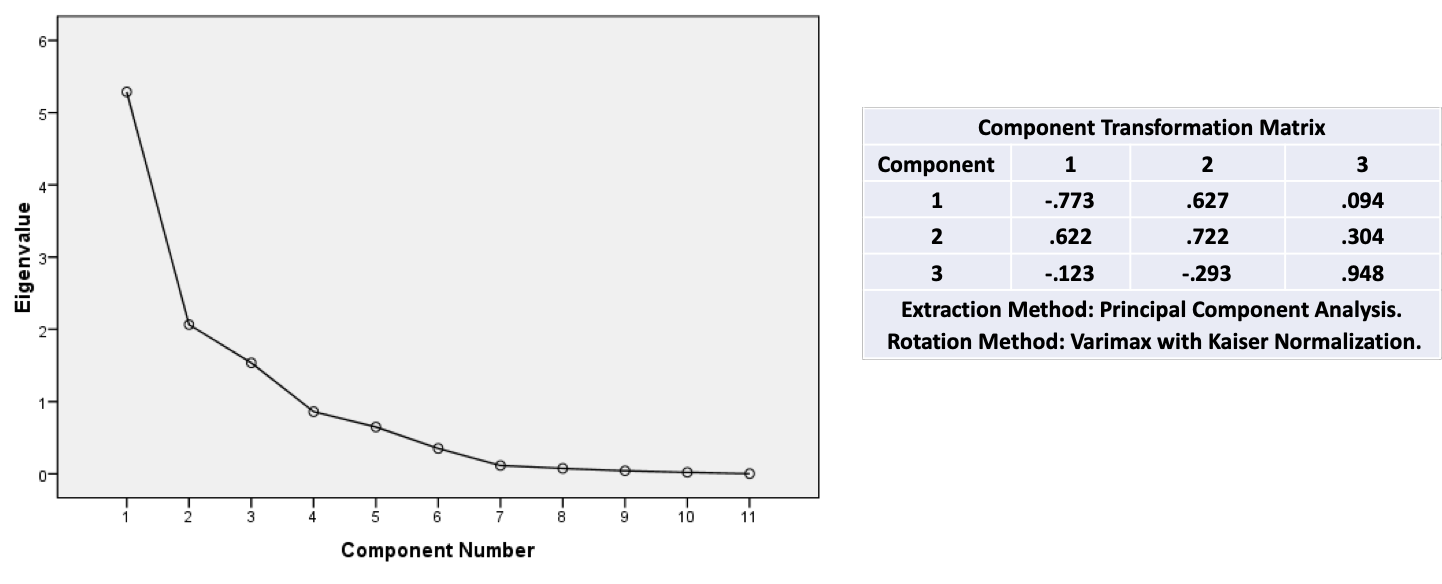

**Table S5: Rotated component matrix with loading for four principle components for treated samples.**

|  | | | |
| --- | --- | --- | --- |
|  | **Component** | | |
| **Parameters** | **1** | **2** | **3** |
| Confirmed | -.466 | .391 | .064 |
| N gene | .888 | -.305 | -.002 |
| ORF gene | .920 | -.128 | -.012 |
| S gene | .946 | -.183 | -.067 |
| Genome | .978 | -.169 | -.029 |
| pH | .157 | .529 | .639 |
| EC | -.322 | .906 | -.048 |
| DO | -.044 | .128 | .783 |
| TDS | -.253 | .943 | .036 |
| ORP | -.124 | -.248 | .762 |
| Salinity | -.265 | .946 | .056 |
| Extraction Method: Principal Component Analysis.  Rotation Method: Varimax with Kaiser Normalization. | | | |
| a. Rotation converged in 5 iterations. | | | |
